# Relative timing of RSV epidemics in summer 2021 across the US was similar to a typical RSV season

**DOI:** 10.1101/2021.09.22.21256632

**Authors:** Zhe Zheng, Joshua L. Warren, Iris Artin, Virginia E. Pitzer, Daniel M. Weinberger

## Abstract

We evaluated the timing of respiratory syncytial virus (RSV) epidemics in 2021 using Google search data. Despite the unusual out-of-season timing, the relative timing of RSV epidemics between states in 2021 shared a similar spatial pattern with typical RSV seasons. These insights can inform prophylaxis administration and planning of clinical trials.

## Introduction

Respiratory syncytial virus (RSV) infection is the most common cause of respiratory hospitalizations in young children [1]. Anticipating the timing of RSV epidemics is important. Administration of available prophylaxis against RSV for high-risk infants needs to be timed with the onset of RSV epidemics. Moreover, the resumption of clinical trials for various RSV prevention strategies needs to be timed to the initiation of the epidemics to ensure sufficient power.

Anticipating epidemic timing for RSV is currently a challenge because the typical seasonal dynamics of the virus were disrupted by restrictions related to the COVID-19 pandemic. In the United States (US), the timing of typical RSV epidemics follows a notable spatial pattern, with epidemics that start earliest in Florida and proceed towards the north and the west [2]. During normal RSV seasons, the epidemics generally begin in the fall and peak in the winter. However, many states in the US observed an unusual increase in RSV cases during the spring and summer of 2021 after the absence of RSV epidemics in the winter of 2020-2021 [3]. As climate conditions differ from typical RSV seasons, these out-of-season RSV epidemics offer an opportunity to probe the drivers of RSV spatial transmission and improve predictions for future seasons.

Google search data have been validated as a good indicator of RSV epidemic patterns [4–6]. It is especially valuable for its timeliness and extensive geographical coverage compared with traditional clinical and laboratory data. Thus, we employed search engine data to estimate the onset of RSV epidemics in each of the 47 continental US states relative to Florida. We set out to evaluate the spatial variation in epidemic timing in summer 2021 compared to a typical season across the US. Our findings can inform the timing of administration of RSV prophylaxis and the planning of clinical trials for RSV vaccines during future epidemics in the US.

## Methods

### Data source on RSV seasonality

We used Google searches as indicators of RSV activity. Google searches for the keyword “RSV” were extracted from Google Trends for the 48 continental states in the US from July 2016 to August 2021 using the R package gTrendsR [7]. Google Trends provides normalized search data by week, which range from 0 to 100, to reflect the relative interests in relevant topics. The timing of search interest for RSV during a typical season and in 2021 is highly correlated with the timing of RSV estimated from the CDC’s National Respiratory and Enteric Virus Surveillance System [4, 6], RSV hospitalizations [4, 6], and pediatric RSV encounters [5].

### Estimating Epidemic Onset at the State Level

We estimated the onset of RSV epidemics for each state in each epidemiological year. The epidemiological year was defined as July to the following June for typical RSV seasons and September 2020 to August 2021 for the out-of-season RSV epidemics. To estimate onset, we fit smooth curves to the search data using generalized additive models with penalized basis splines.

We defined the onset of typical RSV seasons as the time when the second derivative of the smooth functions reached its maximum, implying maximum increase in growth rates of RSV epidemics. We averaged the timing estimates from 2016-2019 and treated this as the onset timing in a typical RSV season.

The second derivative method is less preferable in 2021 as many states have not reached the turning point of the epidemic cycles. It is possible that the increase will continue and the epidemics have not reached the maximum increase in growth rates. Therefore, we determined a threshold based on the average search volume at the onset time in each previous season, with a separate threshold for each state. We then estimated when the search volume crossed these thresholds to determine the epidemic onset time for 2021.

We calculated the relative timing of RSV onset by subtracting the week of RSV onset in Florida (the earliest state) from the week of RSV onset in other states for each season.

### Comparing the relative timing of RSV onset

We used a hierarchical Bayesian regression model to assess the similarity between timing of RSV epidemic onset in a typical year and the timing observed in 2021 across all states (Supplementary Methods). Onset time in a typical year was the predictor, and onset in 2021 was the outcome. We anticipated that if there were no major differences in timing between 2021 and previous years, onset time in a typical year would provide an unbiased estimate of the onset time in 2021(i.e., the intercept would be near zero and the slope would be near one). We included three state-level covariates that could potentially explain differences in the relative timing of RSV epidemics between 2021 and previous years [8]: population density, average household size, and stringency index of non-pharmaceutical interventions against COVID-19 [9]. Additionally, we also included spatially correlated random effects to account for geographic similarity in timing across states. The model is described in detail in the Supplementary Methods. Model parameters and their interpretations are described in Table S1. Data and code to reproduce this study are available from Github: https://github.com/weinbergerlab/RSVtiming.git

## Results

In spring and summer 2021, RSV epidemics started in Florida and proceeded towards the north and west (Figure 1A). States in the northeastern corner (e.g. Maine and Vermont) and the central mountain states had the latest RSV epidemic onsets (Figure 1A).

**Figure 1.**
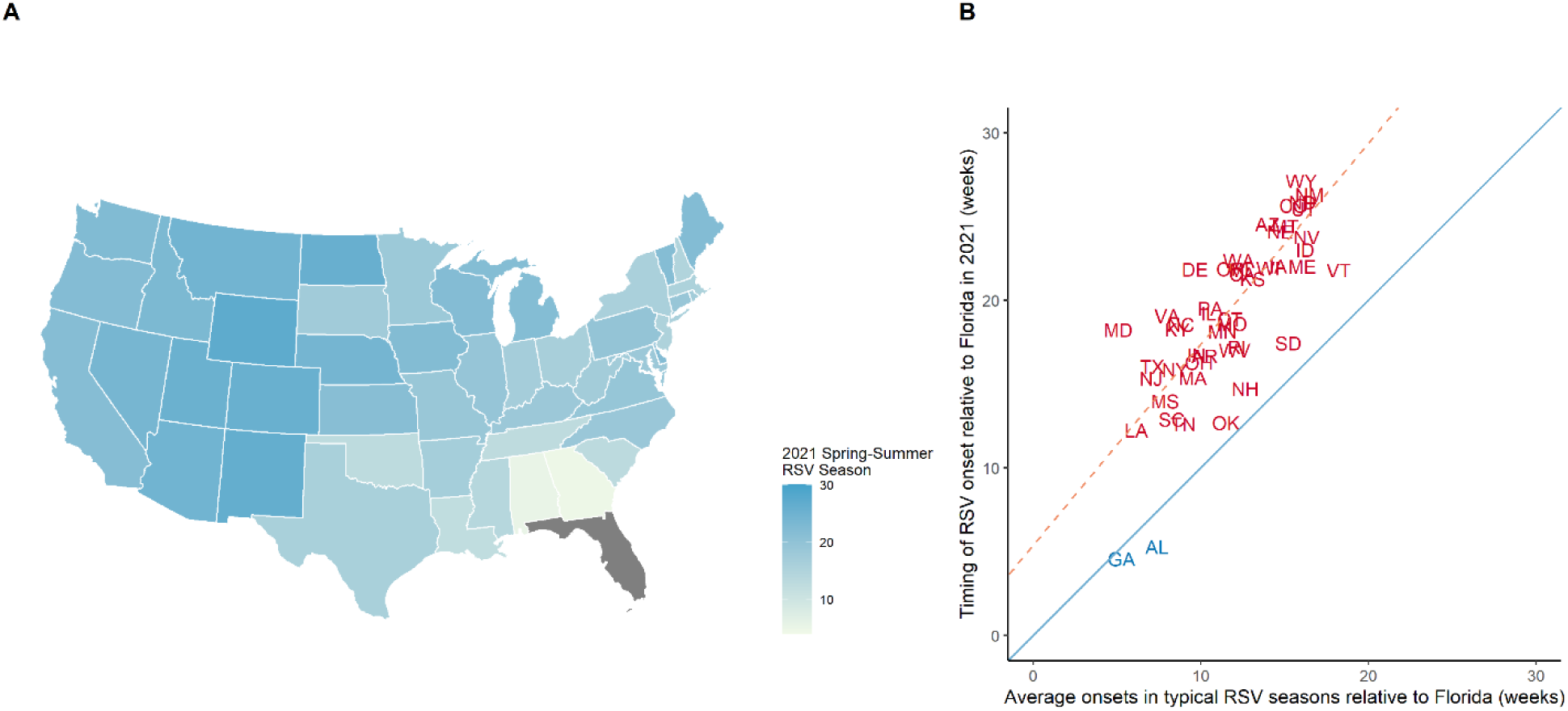
Relative timing of out-of-season RSV epidemics in 2021 across 47 continental US states. A) The map color scale indicates the number of weeks between RSV epidemic onset in 2021 in the state compared to Florida (dark grey); darker colors indicate greater differences in relative timing of RSV onset. B) The scatter plot shows the timing of RSV epidemic onsets relative to Florida in spring-summer 2021 (y-axis) versus the average onsets in typical RSV seasons across 47 US states relative to Florida (x-axis). The solid blue diagonal line is the y=x line; if the relative timing is similar between a typical RSV season and the 2021 RSV season, the states would fall on or near the diagonal line. The dashed orange line is the regression line, showing the estimated relationship across all states.

In general, the relative timing of RSV epidemics in 2021 was 5.4 weeks later relative to Florida compared with that of typical RSV seasons (Figure 1B and Table S1), while the ordering of epidemic onset and speed of spread were similar. Neither the stringency of non-pharmaceutical interventions, population density, nor household size were associated with the shift in the relative timing of the out-of-season RSV epidemics (Table S1). Most states had a delayed timing compared with typical RSV seasons (95% credible intervals for intercepts greater than zero) except for Georgia and Alabama, the two southeastern states that border Florida (Table S1). These states had similar timing relative to Florida for typical RSV seasons and also formed a spatial cluster that was notably different from other parts of the US (Table S1).

## Discussion

Understanding the timing of RSV epidemics is crucial for clinical practice, including administration of prophylactic antibodies against RSV and for planning clinical trials. We sought to better understand the spatiotemporal pattern of RSV epidemics and the underlying factors associated with these patterns. We found that the spatiotemporal pattern of out-of-season RSV epidemics in 2021 was similar to that of typical seasons, but the relative timing of out-of-season RSV epidemics was delayed in most of states compared to three states in the southeastern US. Stringency of non-pharmaceutical interventions, household size, and population density did not explain differences in the relative timing of RSV epidemics in 2021.

Other than two southeastern states (Georgia and Alabama), most states had a delayed start compared to Florida, but similar relative timing compared to previous RSV seasons. Thus, Florida and other southeastern states could serve as a bellwether in future years to adjust timing of the initialization of seasonal prophylaxis administration. Those conducting clinical studies could time recruitment of participants based on the relative timing in other states.

The reason for the delayed timing of RSV onset in other states compared to Florida, Georgia, and Alabama is unclear. However, the onset of the out-of-season RSV epidemics in other states coincided with increased volume of domestic air travel [10]. Future genomic research on virus circulation may help to explain whether these dynamics result from regional and national patterns of movement and transmission or whether other local factors are responsible.

Previous studies have suggested that climate factors play an important role in RSV seasonality and spatiotemporal patterns [11]. However, given that the spatiotemporal pattern of RSV epidemics in spring-summer 2021 was similar to that of previous fall-winter epidemics, it is unlikely that climate factors alone can explain variations in epidemic timing between locations.

Our study is subject to several limitations. First, we used Google search data on RSV as an indicator of RSV activity, which have lower specificity and sensitivity compared with inpatient and laboratory surveillance. While several studies validated the correlations between Google search data and clinical data sources [4, 5], these correlations could be disrupted by increased attention to RSV in the news media, which can influence search behavior. Second, our analysis was performed at the state level, which neglected potential local variations. The relatively small number of data points may also lead to wide confidence intervals, masking the significance of potential predictors of relative timing. Future studies using finer spatial units may help improve the accuracy of projections.

In conclusion, the unusual out-of-season RSV epidemics in 2021 followed a similar spatiotemporal pattern compared to typical RSV seasons. The onset of RSV epidemics in Florida can serve as a baseline to adjust the initiation of prophylaxis administration and clinical trials in other states.

## Author contributions

ZZ performed the analyses and drafted the article. JLW designed the study’s analytic strategy and helped prepare the Methods sections of the text. IA provided the R tool used in the analyses. VEP consulted on the analyses and revisions of the manuscript. DMW conceptualized and designed the study, and reviewed and revised the manuscript. All authors have seen and approved the final draft of the manuscript.

## Supporting information

Supplementary methods and results

## Data Availability

Data and code to reproduce this study are available from Github: https://github.com/weinbergerlab/RSVtiming.git

