## Supplementary methods and results for "Relative timing of RSV epidemics in summer 2021 across the US was similar to a typical RSV season"

**SUPPLEMENTARY TEXT**

**Supplementary Methods**

**Supplementary Results**

**Supplemental Methods:**

We used resampling techniques to obtain confidence intervals for onset timing, using the pspline.inference package in R [1]. For each year, we obtained 150 onset estimates for each state. We averaged a total 600 onset estimates for each state from 2016-2019 and treated this as the onset timing in a typical RSV season. Likewise, the timing estimates in each state in 2021 were the average of 150 samples. We calculated the relative timing of RSV onset by subtracting the week of RSV onset in Florida (the earliest state) from the week of RSV onset in other states for each season.

We used a hierarchical Bayesian regression model to determine if the timing of RSV epidemic onset relative to Florida (the earliest state) in 2021 was similar to the relative timing from a typical RSV season. We also included state-level covariates and spatially correlated random effects to explain potential differences in relative timing in 2021 and to account for geographic similarity in timing across the states, respectively. The model is given as:

$Y_{i}=\alpha_{0i}+\beta_{1} \overline{Z}_{i}+\epsilon_{i},$ and

$$\alpha_{0i}=\beta_{0}+{\mathbf{x}_{i}}^{T}\boldsymbol{\lambda}+\theta_{i}$$

where $Y_{i}$ is the relative timing of RSV epidemic onset with respect to Florida in 2021 in state *i* (measured in weeks), and $\overline{Z_{i}}$ is the average relative timing of RSV seasons in state *i* compared to Florida from 2016-2019. The global intercept parameter $\beta_{0}$ and slope parameter $\beta_{1}$ describe the overall similarity between a typical year’s relative onset timing across all states and the relative timings observed in 2021. The observation-level random effect $\epsilon_{i} \sim N(0, \sigma^{2})$ accounts for residual variability in the data.

The state-specific intercepts, $\alpha_{0i}=\beta_{0}+{\mathbf{x}_{i}}^{T}\boldsymbol{\lambda}+\theta_{i}$, represent the state-level similarity in relative timing for 2021 compared to previous seasons, where $\mathbf{x}_{i}$is the vector of covariates that could potentially explain differences in the relative timing of RSV between 2021 and previous years. The covariates include population density, average household size, and stringency index of non-pharmaceutical interventions against COVID-19.

Additionally, we included a spatially correlated random effect $\left( \theta_{i} \right)$ to adjust for spatial variability in the data in order to ensure accurate inference for the regression parameters of interest. Mapping these estimated parameters can also provide insight into remaining geographic variability in timing that is left unexplained by the included covariates. The spatial effects are assigned the Leroux version of the conditionally autoregressive prior distribution [2], such that

$$\theta_{i}|\boldsymbol{\theta}_{-i},\rho, \tau^{2} \sim\mathrm{MVN}\left( \frac{\rho\sum_{j=1}^{n} w_{ij}\theta_{i}}{\rho\sum_{j=1}^{n} w_{ij}+1-\rho},\frac{\tau^{2}}{\rho\sum_{j=1}^{n} w_{ij}+1-\rho} \right),$$

where $\rho\in[0,1)$ represents the spatial correlation that was estimated from the data; $\rho=0$ corresponds to spatial independence, simplifying the model to an independent random effects model, and 𝜌 near 1 indicates strong spatial correlation. The $w_{ij}$ are variables that contain information about spatial proximity between two locations; $w_{ij}$ equals one if regions *i* and *j* share a common border and equal zero otherwise ($W_{ii}=0$ for all *i*). The total variance of spatial random effects is described by $\tau^{2}$.

We used weakly informative prior distributions to complete the model specification as follows:

$\sigma^{2} \sim$ Inverse-Gamma (0.01,0.01)

$$\beta_{j} \sim N\left( 0, {100}^{2} \right), j=0,1$$

$$\lambda_{j} \sim N\left( 0, {100}^{2} \right),j=1,2,3$$

$$\rho\sim U\left( 0,1 \right)$$

$\tau^{2} \sim$Inverse-Gamma (0.01,0.01).

We fit the model using a Markov chain Monte Carlo posterior sampling algorithmin R [3]. We collect 10,000 samples after removing 100,000 burn-in iterations and thinning the remaining 100,000 posteriors samples by a factor of 10. Convergence was assessed by examining individual parameter trace plots, with no obvious signs of non-convergence observed.

**Supplementary Results**

**Table S1. Interpretation of the differences and similarities in the relative timing of RSV epidemics between typical RSV seasons and spring-summer 2021.**

| Variable | Interpretation | Mean | 95% CrI |
| --- | --- | --- | --- |
| $\beta_{0}$ | $\beta_{0}$ represents the average shift in relative timing (in weeks) between 2021 and typical RSV seasons across states. It would be 0 if there was no difference; values greater than 0 represent later relative timing compared to previous seasons, whereas values less than 0 represent earlier relative timing. | 5.4 | (1.0, 10.2) |
| $\beta_{1}$ | $\beta_{1}$ represents the ordering of epidemic onset and speed of epidemic spread in 2021 compared with typical seasons across states. It would be 1 if there was no difference; values greater than 1 represent slower spatial spread (i.e. greater relative state-to-state differences) compared to previous years, while values less than 1 represent faster spread. | 1.2 | (0.7, 1.5) |
| $\lambda_{1}$ | $\lambda_{1}$, $\lambda_{2}$, and $\lambda_{3}$ describe the associations between relative epidemic timing and average household size, population density, and stringency of non-pharmaceutical interventions, respectively. They would be 0 if there was no association. | 0.4 | (-0.9, 1.7) |
| $\lambda_{2}$ |  | 0.4 | (-0.7, 1.5) |
| $\lambda_{3}$ |  | 0.9 | (-0.1, 1.8) |
| $\alpha_{0GA}$ | $\alpha_{0i}$ represents the state-specific shift in relative timing (in weeks) between 2021 and previous RSV seasons for state *i*. If the relative timing (compared to Florida) is the same in 2021 as compared to previous RSV seasons, $\alpha_{0i}$ would not be significantly different from 0. | -0.4 | (-4.4, 5.3) |
| $\alpha_{0AL}$ |  | 0.7 | (-3.6, 7.0) |
| $\alpha_{0others}$ |  | Lower bound greater than 0 | |
